# Epidemiological description and analysis of *RdRp, E* and *N* genes dynamic by RT-PCR of SARS-CoV-2 in Moroccan population: Experience of the National Reference Laboratory (LNR)-UM6SS

**DOI:** 10.1101/2020.06.18.20135137

**Authors:** Houda Benrahma, Diawara Idrissa, Smyej Imane, Rahoui Jalila, Meskaouni Nida, Benmessaoud Rachid, Arouro Khadija, Jaras Khadija, Moujid Fatima Zahra, Adam Zahra, Nahir Salma, Aouzal Zineb, Elguezzar Hajar, Jeddane Leila, Ousti Fadoua, EL Bakkouri Jalila, Nejjari Chakib

## Abstract

The coronavirus disease 2019 (COVID-19), caused by severe acute respiratory syndrome coronavirus 2 (SARS-CoV-2), is a new infectious disease that first emerged in Hubei province, China, in December 2019. On 2 March 2020, the Moroccan Ministry of Health confirmed the first COVID-19 case in Morocco. The new virus SARS-CoV-2 was identified in the sample of a Moroccan expatriate residing in Italy. Without a therapeutic vaccine or specific antiviral drugs, early detection and isolation become essential against novel Coronavirus.

This study aims to analyze the epidemiological profile of the SARS-CoV-2 in Moroccan cases and to investigate the dynamic of *RdRp, N*, and *E* genes in patients from diagnosis until the recovery.

Among 859 COVID-19 RT-PCR tests realized for 376 patients, 187 cases had positive results COVID-19. 4% were positive with the 3 genes *RdRp, N*, and *E*, 40 % with *N* and *E* genes, 3% with *RdRp* and *N* genes, 31% with only the *RdRp* gene and 22% cases are positives with *N* gene. The analysis of the Covid-19 genes (*RdRp, N*, and *E*) dynamic reveal that more than 6% stay positive with detection of the *N* and *E* gene, and 14% with the *N* gene after 12 days of treatment.

The median period from positive to the first negative Covid-19 RT-PCR tests was 6.8±2.24 days for 44% cases, 14.31± 2.4 days for 30%, and 22.67 ± 1.21 days for 4%.

This a first description of the Moroccan COVID-19 cases and the analysis of the dynamic of the *RdRp, N*, and *E* genes. The analysis of our population can help to improve in the care of patients.

## 1. Introduction

According to the World Health Organization (WHO), the WHO China Country Office was informed of cases of pneumonia of unknown etiology in Wuhan City, Hubei Province, on December 31, 2019 [1].

The investigations identified a new virus that was closely related to severe acute respiratory syndrome coronavirus (SARS -CoV) [2].The novel coronavirus in humans was initially named as 2019-nCoV [3], and then designated as SARS-CoV-2 by the Coronavirus Study Group of the International Committee on Taxonomy of Viruses [4]. And WHO announced the epidemic disease caused by SARS-CoV-2 as coronavirus disease 2019 (COVID-19) [5].

On April 28^th^, 2020, 213 countries and territories included Morocco, with more than 2 959 929 confirmed cases and more than 202 733 have died from the rapidly-spreading SARS-CoV-2 virus [6].

On the 2sd of March 2020, the Ministry of Health confirmed the first COVID-19 case in Morocco. The virus was detected in a Moroccan expatriate residing in Italy and who came from Italy on February 27^th^, 2020. Also, the second case was confirmed by the end of the same day, involving an 89-year-old woman Moroccan residing in Italy too who had returned to Morocco on the 25^th^ of February 2020. Till the 28^th^ of April 2020, 4,246 confirmed cases of COVID-19 with 163 deaths [7].

Among the foremost priorities to facilitate public health interventions in Morocco is a reliable laboratory diagnosis. For that, the Ministry of Health expands screening tests across the country as part of the country’s preparation to end confinement measurement and to rapidly evaluate more possible cases. On April 1^st^, 2020, the National Reference Laboratory (LNR) of Mohammed VI University of Health Sciences (UM6SS), became the fourth laboratory in Morocco authorized to carry out the biological screening and diagnostics of COVID-19.

In acute respiratory infection, RT-PCR is routinely used to detect causative viruses from respiratory secretions. For ensuring the diagnostic of Covid-19, the LNR has been equipped with different platforms to performed real-time RT-PCR testing for three targets in the virus: the envelope (E), the RNA dependent RNA polymerase (RdRp) and the nucleocapsid (N).

The coronavirus SARS-CoV-2 genome consists of a leader sequence, ORF1ab encoding proteins for RNA replication, and genes for non-structural proteins (nps) and structural proteins [8]. Like other betacoronaviruses, the SARS-CoV-2 genome encodes four major structural proteins. The structural proteins are involved in various viral processes, including virus particle formation. The structural proteins include spike (S), envelope (E), membrane protein (M), and nucleoprotein (N), which are common to all coronaviruses [9, 10].

To date, no studies exploring the variation of *RdRp, N* and *E* genes expression of SARS-CoV-2 in the patient’s specimen. In this study, in first time, we analyses the epidemiological profile of the SARS-CoV-2 in Moroccan cases and in second time we explored the dynamic of *RdRp, N* and *E* genes in patients from diagnosis until the recovery.

## 2. Material and Methods

### 2.1 Patients and samples

All samples included in this study were routinely tested for the presence of SARS-CoV-2 immediately upon arrival in the National Reference Laboratory (LNR) from the Cheikh Khalifa International University Hospital, Mohammed VI University of Health Sciences (UM6SS). In this study, we included a total of 376 patients admitted to hospital between March 28, 2020, and April 29, 2020.

Nasopharyngeal swab samples were collected for extracting SARS-CoV-2 RNA from patients suspected of having COVID-19 infection. The collected swabs were placed into the transport tube where includes a universal transporting medium that is room temperature stable (UTM-RT).

The LNR provide a RT-PCR to clinically suspected COVID-19 patients when they were admitted. Multiple repeat RT-PCR tests were performed during admission in the hospital in a different period based on national mandatory management guidelines.

### 2.2. RNA extraction

The RNA extraction from nasopharyngeal samples was performed using AccuPrep ® Viral RNA Extraction Kit (Bioneer Corporation, Korea) according to the manufacturer’s instructions. Briefly, swabs were vortexed for 10 second speed followed by extraction from 200 µl of UTM-RT, and all samples were subjected to extraction with an elution volume 50 μl.

### 2.3. RT-PCR SARS-CoV2 detection

One-Step Reverse Transcription Real-Time polymerase chain reaction (RT-PCR) was used to confirm the presence of the SARS-CoV-2 genome by amplification of *RdRp*, E, and N gene. For the samples included in this study, we realized the RT-PCR by using GeneFinderTM COVID-19 Plus RealAmp Kit according to the manufacturer’s protocol (OSANG Healthcare Co., Ltd, South Korea).

The RT-PCR assays were performed on the CFX96 Touch Real-Time PCR Detection System (Bio-Rad Laboratories, Inc.). The reaction mixture contains 10 μl of COVID-19 PLUS Reaction Mixture, 5μl of COVID-19 PLUS Probe Mixture, and 5μl of sample RNA. Total Reaction volume is 20μl per sample.

The RT-PCR assays were performed under the following conditions: reverse transcriptional reaction at 50 °C for 20 minutes, pre-denaturation at 95 °C for 5 minutes, 45 cycles of denaturation at 95 °C for 15 seconds and extending and collecting fluorescence signal at 58 °C for 60 seconds.

The result interpretation was performed according to the manufacturer’s protocol (table 1)

**Table 1:**
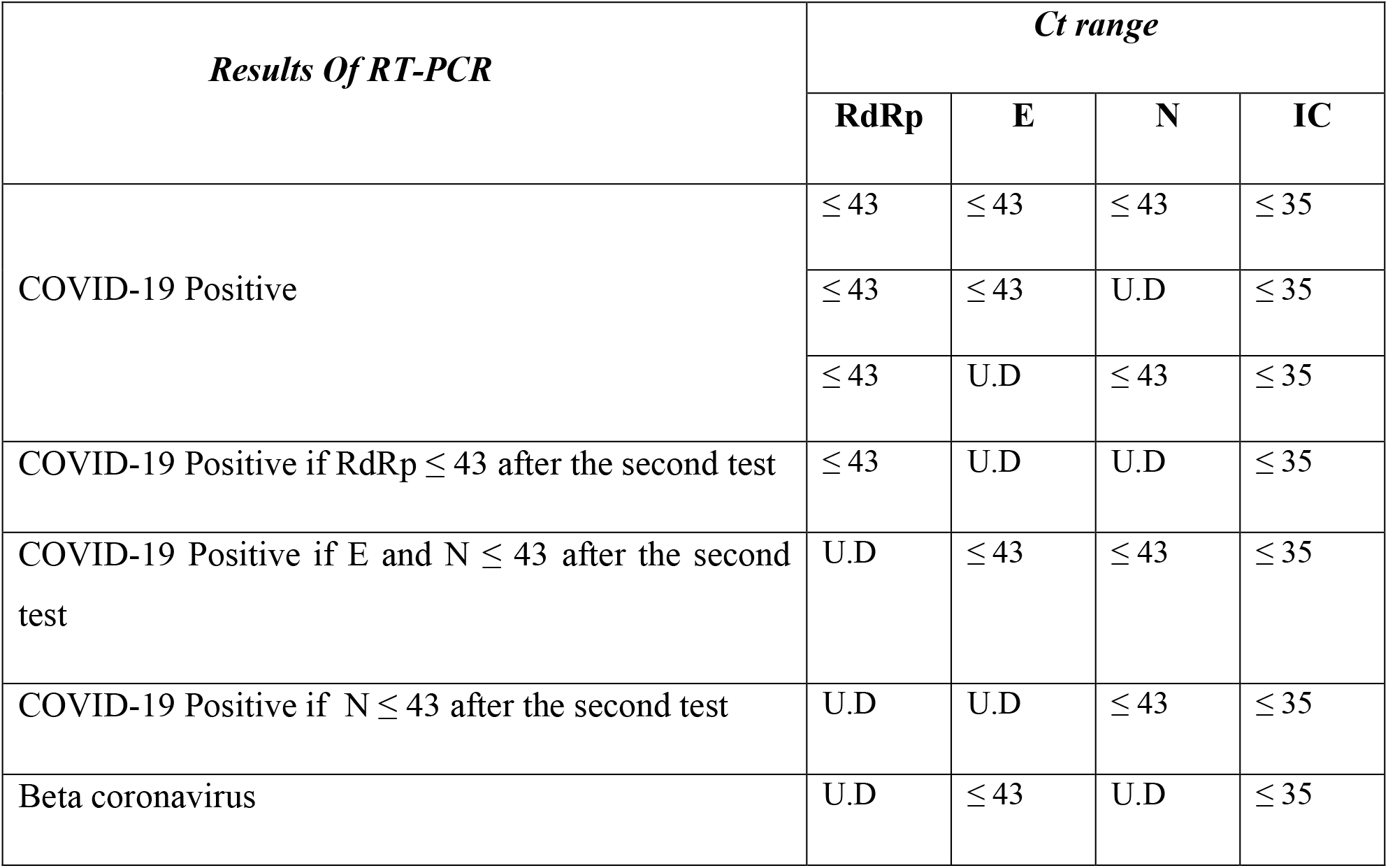
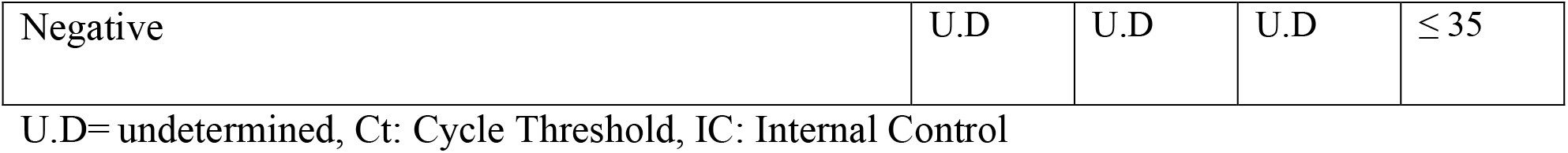
Results interpretation of GeneFinder™ COVID-19 Plus RealAmp Kit

| *Results Of RT-PCR* | *Ct range* |  |  |  |
| --- | --- | --- | --- | --- |
|  | **RdRp** | **E** | **N** | **IC** |
| COVID-19 Positive | ≤ 43 | ≤ 43 | ≤ 43 | ≤ 35 |
|  | ≤ 43 | ≤ 43 | U.D | ≤ 35 |
|  | ≤ 43 | U.D | ≤ 43 | ≤ 35 |
| COVID-19 Positive if RdRp ≤ 43 after the second test | ≤ 43 | U.D | U.D | ≤ 35 |
| COVID-19 Positive if E and N ≤ 43 after the second test | U.D | ≤ 43 | ≤ 43 | ≤ 35 |
| COVID-19 Positive if N ≤ 43 after the second test | U.D | U.D | ≤ 43 | ≤ 35 |
| Beta coronavirus | U.D | ≤ 43 | U.D | ≤ 35 |
| Negative | U.D | U.D | U.D | $\leq 35$ |

U.D= undetermined, Ct: Cycle Threshold, IC: Internal Control

### 2.4. Statistical analysis

Continuous variables were presented as means ± standard deviation (SD). For categorical variables were presented as counts and percentages. P-values less than 0.05 were considered as statistically significant. All statistical analyses were performed using STATA software, version 11.0.

### 3. Results

### 3.1. General characteristics

In this study, the total number of Covid-19 RT-PCR assays realized on the 376 included COVID-19 patients was 859, with 3.7 tests per patient. The mean age was 46.82 ± 20.54 years old, comprising 55.76 % men and 44.24% women (table 2).

**Table 2:**
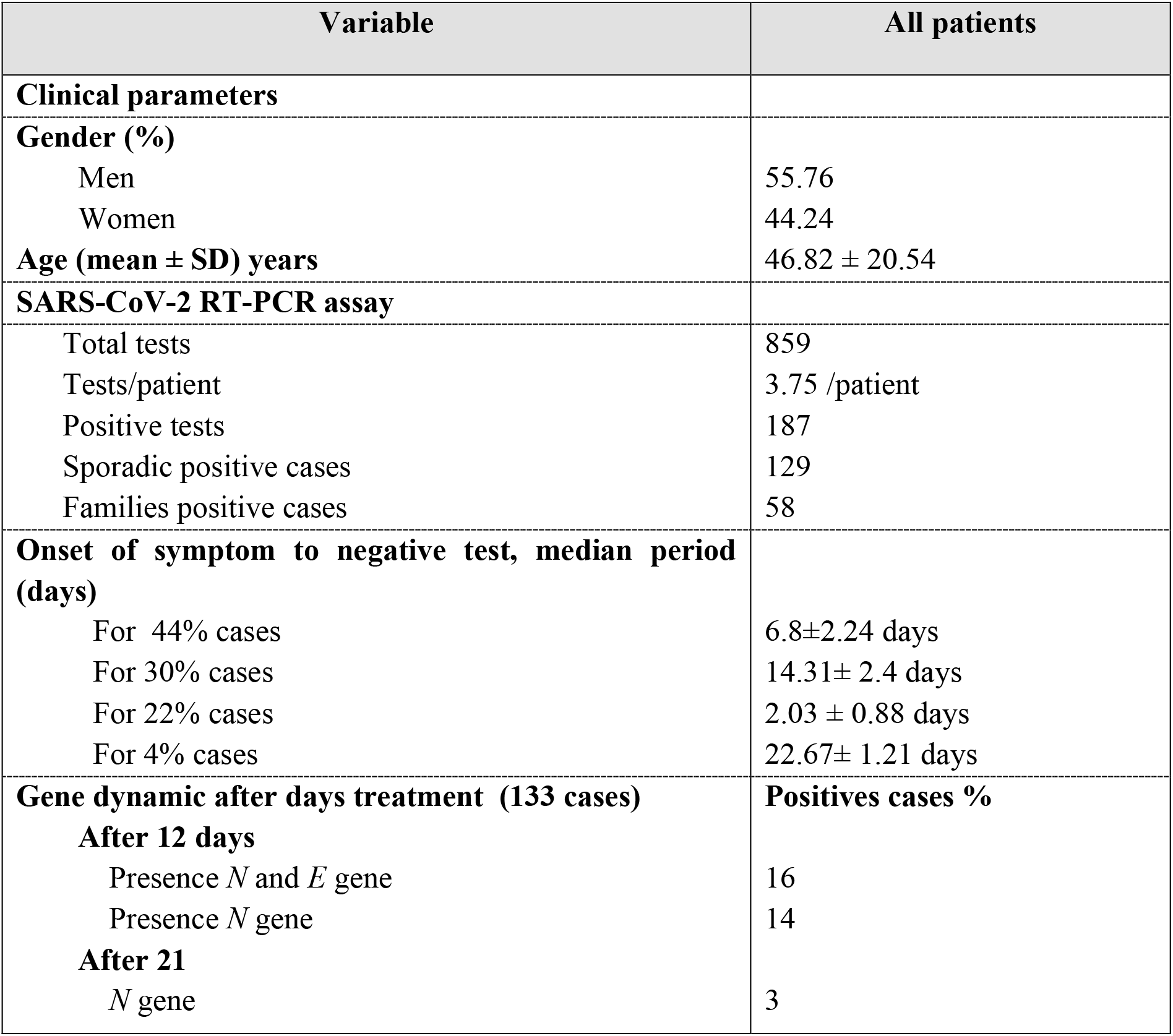
clinical characteristics of all patients, and gene dynamic in positives cases

| Variable | All patients |
| --- | --- |
| **Clinical parameters** |  |
| **Gender (%)** |  |
| Men | 55.76 |
| Women | 44.24 |
| **Age (mean ± SD) years** | $46.82 \pm 20.54$ |
| **SARS-CoV-2 RT-PCR assay** |  |
| Total tests | 859 |
| Tests/patient | 3.75 /patient |
| Positive tests | 187 |
| Sporadic positive cases | 129 |
| Families positive cases | 58 |
| **Onset of symptom to negative test, median period (days)** |  |
| For 44% cases | $6.8 \pm 2.24$ days |
| For 30% cases | $14.31 \pm 2.4$ days |
| For 22% cases | $2.03 \pm 0.88$ days |
| For 4% cases | $22.67 \pm 1.21$ days |
| **Gene dynamic after days treatment (133 cases)** | **Positives cases %** |
| **After 12 days** |  |
| Presence *N* and *E* gene | 16 |
| Presence *N* gene | 14 |
| **After 21** |  |
| *N* gene | 3 |

Of the 376 patients, 189 (50.3 %) had negative results and 187 (49.7%) had positive results for Covid-19 RT-PCR tests. In the negative cases, the men present 53.16%, and the women 46.84% (table 2).

In the Covid-19 positives cases, the gender distribution is 41.83% women and 58.14% men.

Among the Covid-19 positive patients, we identify 129 sporadic and 58 family cases (with known exposure history). Family with SARS-CoV-2 infection corresponds to 24 families. The analysis of these families showed that 71% included 2 cases, 21% included 3 cases, 4% included 4 cases, and 4% included 5 cases,

### 3.2. SARS-CoV-2 Genes dynamic

The interpretation of the RT-PCR results for the first diagnosis of the three genes *RdRp, N*, and *E* showed that: 4% of cases are positive with the 3 genes *RdRp, N*, and *E*. 31% cases are positive with only the *RdRp* gene, 3% of cases are positive with *RdRp* and *N* gene, 22% cases are positive with *N* gene, and 40 % with *N* and *E* gene. (tab2)

The analysis of the Covid-19 genes *RdRp, N*, and *E* dynamic revealed that more than 6% of the SARS-CoV-2 infected person stay positive with detection of the *N* and *E* gene, and 14% of the positive patients stay positive with the *N* gene after 12 days the treatment. After more than 21 days of treatment, the RT-PCR test reveals the persistence of the *N* gene in 4 cases (3%).

From symptoms and positive Covid-19 RT-PCR test to the first negative Covid-19 RT-PCR tests, the median period was 6.8±2.24 days for 44% cases.

For 30% cases the median period 14.31± 2.4 days for. A longer period was identified in 4% of cases with a median period of 22.67 ± 1.21 days. For 22% of cases the median period was 2.03 ± 0.88 days, for these patients the RT-PCR was realized for the first time by other reference laboratories (tab2). For all patients with the first negative test for the 3 genes, a second RT-PCR test was realized after 24h (table 2).

We also investigate the impact of age on the profile of Covid-19 by RT-PCR. The Test of homogeneity is carried out by comparing the positive and negative cases for a different range of age. A significant association was observed for the over than 64 years old range with p = 0.009 and OR= 1.74, 95% CI= [1.28, 2.33].

## 4. Discussion

The current SARS-CoV-2 outbreak is the third epidemic attributed to coronavirus in the 21st century, and incredibly the number of confirmed SARS-CoV-2 infection has surpassed SARS and MERS in world wild [1, 11].

This is the first scale report from 285 COVID-19 Moroccan patients with 859 samples of RT-PCR tests for Covid-19 detection. The data of this study is the preliminary analysis of the Moroccan population analyzed in the LNR which will allow making more in-depth studies of patients infected by the SARS-CoV-2.

After analysis of the cases implicated in our study, we found that the infection was prone to affect mem more than women with different percentages of 55.76% and 44.24% respectively. This result consistent with many conclusions from different studies [12, 13].

Many studies suggested that coronavirus infected older more than young individuals [14]. In our study, comparing positive and negative cases for a different range of age we found a significate difference (p = 0.009 and OR= 1.74, 95% CI= [1.28, 2.33]), for the range age ≥ 64 years. Various studies confirm our observation which suggests that older individuals are more likely to be infected by the virus [12, 15]. We can explain the correlation between age and virus by the presence of higher levels of angiotensin converting enzyme 2 in older people alveoli which are thought to be a receptor for SARS [16].

This study included 129 sporadic and 58 familial cases. Various studies analyses the mechanism of transmission of SARS-CoV-2, and they confirmed that the Human to Human transmission via droplets is the main route of transmission within a susceptible population. But no rule out transmission by asymptomatic carriers [17]. The first cases in Morocco were traveled from the epidemic region in Italy. And family members who traveled from Europe were most likely responsible for a familial cluster of COVID-19 once back home. Another explication of the presence of clusters in our country is the organization of familial activity (wedding, death ceremonies, etc…), and some industrial activity.

The SARS-Cov-2 share similar sequencing characteristics with SARS-CoV and MERS-CoV, but the analysis of different case series showed that the shedding pattern of the viral nucleic acid of patients infected with SARS-CoV-2 is different from SARS-CoV. In the early stage, the SARS-CoV had a modest viral load peaked approximately 10 days after symptoms onset.

For the SARS-CoV-2, the median duration of the virus in the respiratory sample was 18 days, and from symptom onset, the peak viral shedding in respiratory specimens of positive cases occurred after about 10 to days for the SARS-CoV [18, 19]. In our study, we found that the duration of virus shedding in lower respiratory tract samples was longer than 14 days for 30% of cases, and peak viral shedding occurred after about two weeks from symptom onset. Tracing the duration of the virus is very important for effective control and prevention of the epidemic.

Because highly sensitive and specific diagnostic assays are the key to the identification of cases, contact tracing, for diagnosis of cases with suspected SARS-CoV-2 infection. We chose to performed detection of unique viral sequences with RT-PCR by using GeneFinderTM COVID-19 Plus RealAmp Kit (OSANG Healthcare Co., Ltd, South Korea), which is one of the validated protocols for in vitro diagnostic (CE marked) available on the market. The RT-PCR assays target the E gene encoding for the envelope protein, which is common to the Sarbecovirus subgenus, the second specific assay targets the *RdRp* gene encoding for RNA-dependent RNA polymerase and the third assay target the *N* gene encoding nucleocapsid protein [20, 21].

Based on the RT-PCR tests of positive cases during hospitalization period, we found that after 12 days of treatment, more than 6% of the SARS-CoV-2 infected person stay positive with the detection of the *N* and *E* gene, and 14% of the positive patients stay positive with the *N* gene. And only 3% stay positive with the N gene after more than 21 days of treatment.

To our knowledge, the dynamic of the three SARS-CoV-2 genes was not analyzed before. The genome SARS-CoV-2 shares similar sequencing characteristics with SARS-CoV and MERSCoV. Human coronaviruses are positive-sense RNA (30 kb) viruses. Two types of proteins characterize Human coronavirus, structural (Spike (S), Nucleocapsid (N), Matrix (M), and Envelope (E)) and non-structural proteins (nsp1 up to nsp16) including the RNA dependent RNA polymerase (RdRp) (nsp12). The organization of the coronavirus genome is 5′-leader-UTR-replicase-S (Spike)-E (Envelope)-M (Membrane)-N (Nucleocapsid)-3′ UTR-poly (A) tail [22].

If we analyze the Coronavirus life cycle, we found that the initial attachment of the virion to the host cell is initiated by interactions between the S protein and its receptor. Following receptor binding, the virus must next gain access to the host cell cytosol. This is generally accomplished by acid dependent proteolytic cleavage of S protein [23]. The RdRp is a vital enzyme for the life cycle of RNA viruses, because the ability of coronavirus to recombine is tied to the strand switching ability of this protein. Recombination likely plays a prominent role in viral evolution [23]. For the nucleocapsid (N) protein, the primary function is to package the viral RNA genome within the viral envelope into a ribonucleoprotein (RNP) complex called the capsid. Ribonucleocapsid packaging is a fundamental part of viral self-assembly and replication. Additionally, the N-protein of the SARS-CoV-2 affects host cell responses and may serve regulatory roles during its viral life cycle [24].

In our study, we found that the N gene persists in cases sample with Ct ≤ 40 in RT-PCR. To explain this persistence, different studies showed that the coronavirus N protein is abundantly produced within infected cells. And these proteins have multiple functions, including binding to viral RNA to form the ribonucleocapsid and have also been proposed to have roles in virus replication, transcription, and translation [24]. Also, in Humans cells, N proteins have been shown to cause deregulation of the cell-cycle, inhibit the production of interferon and induce apoptosis in serum deprived cells, of all which may have possible pathological consequences [24, 25].

## 5. Conclusion

In summary, this is the first description of the dynamic of *RdRp, N*, and *E* genes of the SARS-CoV-2 virus among COVID-19 positive patients. The study allow us to focus on several parameters involved in the care of patients We highlighted that the comprehension of dynamism of these tree genes is an important parameter in the COVID -19 diagnosis. We plan to include in the second time the clinical parameters and the history of infection, to explain this dynamic of this infection. A comprehensive understanding of COVID-19 will help to control the disease.

## Data Availability

The data are not publicly available

## Data Availability

All data underlying the results are available as part of the article, and no additional source data are required.

## Declaration of Competing Interest

All authors declare that there are no conflicts of interest.

## Notes

### Competing Interest Statement

The authors have declared no competing interest.

### Funding Statement

no funding

### Author Declarations

ethics committee of the Mohammed VI University of Health Sciences (UM6SS) of Morocco

